# Genomic Profiling Identifies Somatic Mutations Predicting Thromboembolic Risk in Patients with Solid Tumors

**DOI:** 10.1101/2020.06.09.20124925

**Authors:** Andrew Dunbar, Kelly L. Bolton, Sean M. Devlin, Francisco Sanchez-Vega, Jianjiong Gao, Jodi V. Mones, Jonathan Wills, Daniel Kelly, Mirko Farina, Keith Cordner, Young Park, Sirish Kishore, Krishna Juluru, Neil M. Iyengar, Ross L. Levine, Ahmet Zehir, Wungki Park, Alok A. Khorana, Gerald A. Soff, Simon Mantha

## Abstract

Cancer-associated venous thromboembolism (CAT) is a well-described complication of cancer and a leading cause of death in cancer patients. The purpose of this study was to assess potential associations of molecular signatures with CAT, including tumor-specific mutations and the presence of clonal hematopoiesis. We analyzed deep-coverage targeted DNA-sequencing data of >14,000 solid tumor samples using the MSK-IMPACT™ platform to identify somatic alterations associated with VTE. Endpoint was defined as the first instance of cancer-associated pulmonary embolism and/or proximal/distal lower extremity deep vein thrombosis. Cause-specific Cox proportional hazards regression was used, adjusting for pertinent clinical covariates. Of 11,695 evaluable individuals, 72% had metastatic disease at time of IMPACT. Tumor-specific mutations in *KRAS* (HR=1.34 [1.09-1.64]; adjusted *p*=0.08), *STK11* (HR=2.12 [1.55-2.89]; adjusted *p*<0.001), *KEAP1* (HR=1.84 [1.21-2.79]; adjusted *p*=0.07), *CTNNB1* (HR=1.73 [1.15-2.60]; adjusted *p*=0.09), CDKN2B (HR= 1.45 [1.13-1.85], adjusted p=0.07) and *MET* (HR=1.83 [1.15-2.92]; adjusted *p*=0.09) were associated with a significantly increased risk of CAT independent of tumor type. Mutations in *SETD2* were associated with a decreased risk of CAT (HR=0.35 [0.16-0.79], adjusted p=0.09). The presence of clonal hematopoiesis was not associated with an increased VTE rate. This is the first large-scale analysis to elucidate tumor-specific genomic events associated with CAT. Somatic tumor mutations of STK11, KRAS, CTNNB1, KEAP1, CDKN2B and MET were associated with an increased risk of VTE in solid tumor patients. Further analysis is needed to validate these findings and identify additional molecular signatures unique to individual tumor types.

**Key Points:**

- Tumor mutations in *STK11, KRAS, CTNNB1, KEAP1, CDKN2B, MET* and *SETD2* modulate the risk of cancer-associated thrombosis.
- The presence of clonal hematopoiesis does not affect the risk of cancer-associated thrombosis.

## Introduction

Venous thromboembolism (VTE) is a frequent complication of cancer and a leading cause of cancer-related mortality.^1,2^ Clinical risk factors known to portend an increased risk of cancer-associated venous thromboembolism (CAT) include body mass index (BMI), prior VTE events, inherited thrombophilias, blood cell counts, exposure to chemotherapy or recombinant erythropoietin, cancer stage, and the presence of underlying comorbid conditions, including infection.^3-14^ These factors form the basis of risk assessment models frequently utilized to identify patients most likely to benefit from anticoagulant prophylaxis, most commonly the Khorana score.^3,15^ However, despite improving CAT risk prediction, significant limitations remain with these models, and the optimal approach to pharmacological prophylaxis remains unclear. Identification of additional risk factors to enhance current CAT prediction models might help to better personalize VTE prevention and improve clinical outcomes for solid tumor patients.

The mechanisms driving the pathogenesis of CAT are likely complex and multi-factorial and highlight the inextricably linked and highly dynamic interactions between the tumor, its microenvironment, and the hemostatic system. Known tumor-specific factors shown to promote venous thrombosis in cancer include: 1) over-expression and secretion of various pro-coagulant factors by the tumor, including tissue factor (TF) and TF-bearing microparticles; 2) activation of platelets and/or leukocytes by tumor secreted factors including pro-inflammatory cytokines; and/or 3) secondary effects of tumor cells on surrounding vasculature and tissue microenvironment as a result of aberrant pro-angiogenic growth factor stimulation.^16^ Importantly, VTE rates vary significantly based on tumor sub-type, and even by histological sub-type within tumor types,^17,18^ suggesting that the individual mechanisms promoting thrombosis, and the degree to which the coagulation system is perturbed, are highly tumor-specific.

In this manner, cancer cell genotype is increasingly recognized as an important factor for VTE development.^19^ Tumor-specific mutations in *KRAS*,^20,21^ *ALK*^22,23^ and *ROS1*^24^ have been associated with an increased risk of thromboembolism. Results for *EGFR* mutational status and VTE risk have been mixed, with only one study showing an increased risk and others showing no effect or even a decreased risk of VTE.^20,25-28^ Similarly, while many high-grade gliomas over-express TF in hypoxic conditions, the presence of an *IDH1* mutation appears to be protective against VTE in part due to reduced TF expression, further highlighting the influential role of tumor genotype in influencing CAT risk.^29-32^ Also, evidence from an animal model suggests tumor-specific *MET* mutations might be associated with a hypercoagulable state.^33^ Over-expression of TF and other pro-coagulant factors along with enhanced neoangiogenesis via constitutive VEGF stimulation are proposed mechanisms for the thrombophilic effect of oncogenic mutations.^34^ Whether the pro-coagulant state of these tumors is simply a byproduct of aberrant signaling or serves directly to further promote tumor growth remains unclear; regardless, these data underscore the urgent need to identify the full extent of molecular events that might contribute to cancer-related thrombosis.

The emerging entity of clonal hematopoiesis (CH) is also increasingly recognized to influence thrombotic risk.^35^ CH has been found to occur in >10-20% of otherwise healthy individuals over the age of 70 and in >25% of patients with solid tumor malignancies and is associated with adverse clinical outcomes.^35,36^ Importantly, recent studies have implicated CH in an increased rate of arterial thrombotic events, including myocardial infarction and stroke in otherwise healthy individuals.^37^ In one retrospective series, JAK2 V617F mutant CH was also found to be associated with an increased risk of VTE and early onset atherothrombotic disease;^38^ however, the role of CH influencing rates of VTE within the cancer patient population remains unknown.

While mutational profiling is increasingly used to inform important diagnostic, prognostic, and therapeutic considerations in solid tumor oncology patients, current clinically used assays are limited in the breadth and depth of detectable alterations. Since 2014, a custom hybridization capture-based next-generation sequencing assay known as IMPACT (Integrated Mutation Profiling of Actionable Cancer Targets) has been utilized at Memorial Sloan Kettering Cancer Center (MSKCC) (MSK-IMPACT) to comprehensively profile patient tumors at the molecular level, with the capacity to detect somatic alterations in over 300 genes at a minimum depth of coverage of 91X.^39^ To date, over 40,000 patient tumors have been profiled by MSK-IMPACT.^40^ Given the availability of this platform and the unmet need of improving VTE prediction across the oncologic patient population, we sought to retrospectively analyze solid tumor MSK-IMPACT data to assess molecular determinants of CAT in patients treated at MSKCC.

## Methods

### Patients and data capture

We included all adult patients who had solid tumor MSK-IMPACT testing between January 2014 and December 2016, as well as available tumor registry and electronic medical record (EMR) data **(Appendix A)**. All patient samples and data were obtained following MSKCC institutional review board approval. Cancer-associated VTE was defined as any instance of pulmonary embolism (PE) or lower extremity deep vein thrombosis (DVT), including both proximal and distal DVT events. Symptomatic and asymptomatic VTE episodes were included. Radiology reports from a list of studies susceptible to elicit a diagnosis of PE or lower extremity DVT **(Appendix B)** for the years 2011 to 2016 were searched for keywords indicative of VTE **(Appendix C)**, and all positive reports were reviewed and verified by an observer (SM) for the presence of a VTE episode. The pre-IMPACT period from 2011-2013 was included to ensure that VTE episodes occurring in the years prior to cohort entry were identified. The R statistical platform was used along with package TM for the text mining portion of the work. Parallel to this search, pharmacy records were screened for mentions of therapeutic doses of an anticoagulant **(Appendix D)**, and positive findings were reviewed by an observer (SM) for patients not already noted as a case in the radiology text search. All cases discovered in the radiology and anticoagulation search were reviewed and confirmed by a second observer (AD). Lastly, the corpus of medical notes ranging the years 2013 to 2016 was processed with a customized natural language processing (NLP) pipeline and data entry interface to identify missing lower extremity DVT and PE cases. Only events having occurred up to one year before cancer diagnosis were considered to be cancer-related; older episodes were recorded separately. Upper extremity DVT events were captured using the same customized NLP pipeline with each case reviewed by an observer (JM and SM). Information on tumor type and basic demographic characteristics was obtained from the institutional tumor registry and clinical database system. In order to facilitate the analysis and interpretation of the results, tumor type was simplified as detailed in **Appendix E**. Metastatic status during the study period was estimated by merging staging data from the tumor registry and IMPACT records **(Appendix F)**. Patients not marked as metastatic upon cohort entry had their clinical notes processed with the NLP pipeline to pick up any missed instances of metastatic disease. A manual audit was conducted on 300 randomly selected patients to assess the accuracy of data collection and aggregation.

### MSK-IMPACT Sequencing

The MSK-IMPACT assay was performed as previously described.^39^ Briefly, DNA from both tumors and patient-matched blood samples were obtained. Bar-coded libraries were then generated and sequenced. The custom targeted gene panel consisted of 341 genes **(Appendix G)**. Subsequent versions of MSK-IMPACT contain additional numbers of genes (410 genes in Version 2; 468 in Version 3); however, these were ultimately excluded from the final analysis to maintain consistency across the cohort. Mean coverage across all tumor samples was 753X, with minimum depth of coverage of 91X. Raw results were run through a custom pipeline to identify somatic alterations. Germline mutations were filtered out. Final data from all samples were made publicly available online through the *cBioPortal for Cancer Genomics*.^41,42^ For the current analysis, mutation data for all individuals were mapped as binary values (mutated vs unmutated). Only putative driver mutations were retained. Data on copy number alterations and gene fusions were also evaluated. In the case of fusions, only the gene exhibiting a potential change in expression was considered mutated. Fusions detected with the Archer panel (Illumina) were excluded, even though listed in *cBioPortal*, because only a small number of samples had this assay performed.

### Statistical Analysis

The primary endpoint of time-to-CAT was defined as the time from accession to the minimum of CAT development, upper extremity DVT, death, or last follow-up in a one-year period following accession. CAT included lower extremity DVT (proximal or distal) and pulmonary embolism, whether symptomatic or not. Accession was defined as the date of blood sample receipt for IMPACT testing, and the approximate time a patient consents to testing. Cause-specific Cox proportional hazards regression estimated associations between select somatic mutations and the risk of developing a CAT episode. A separate model was built for each mutation; all models were adjusted for cytotoxic chemotherapy as a time-dependent covariate, history of VTE, current anticoagulation use, presence of metastatic disease, and age at accession. Additionally, all models were stratified based on tumor type, and because some tumor samples were previously banked, the years from the procedure to accession ([0], (0-0.25], (0.25-1], (1-5], [5,+)). Left truncation was used for the subset of procedures that occurred after accession but before the end of the one-year period. We only considered the 50 most commonly mutated genes in the cohort, in addition to *MET, ALK*, and *ROS1* which were selected based on prior data suggesting an effect on the risk of thromboembolism in cancer.^22-24,33^ The Benjamini-Hochberg method was used to adjust *p*-values for false discovery. The predetermined statistical significance cutoff for the purpose of this analysis was 0.10. The R 3.6.1 statistical platform was used for multivariate analysis. For original data, please contact.

## Results

### Patient characteristics and incidence of VTE

The schema used for VTE assessment in all patients is delineated in **Figure 1**. From 2014 to 2016, a total of 14,223 adult patients with solid tumor malignancy had MSK-IMPACT genomic sequencing and met initial inclusion criteria for this retrospective study. Of these, 1,962 were found to have at least one episode of CAT. 2,528 patients were excluded from cohort entry due to pre-existing CAT (n=1,104), upper extremity DVT (n=161) or because they had no clinical notes available in the EMR after the start of observation, consisting of the latest of IMPACT tissue sampling or blood sample accession. A total of 11,695 individuals were included in the final analysis, with 693 episodes of CAT recorded in the first year of observation. The highest event rates were in the sub-group of patients with pancreatic cancer **(Figure 2)**. Characteristics of the patient cohort are presented in **Table 1**. The prevalence of individual cancer types included in the entire IMPACT cohort were generally reflective of the prevalence of cancer types within the larger oncologic patient population; however, breast cancer and prostate cancer, two of the most common cancer types, were under-represented, likely owing to fewer patients with these tumor types undergoing extended genomic testing during the observed time period. 163 patients (1.4%) had a VTE episode documented more than one year prior to cancer diagnosis. Importantly, most patients (72%; N=8,383) had metastatic disease at time of MSK-IMPACT.

**Table 1:** Characteristics of Patients.

|  |  | Overall<br>N (%) | VTE Event<br>N (%) |
| --- | --- | --- | --- |
| **Cytotoxic**<br>**Chemotherapy\*** | **No** | 6759 (58%) | 402 (58%) |
|  | **Yes** | 4936 (42%) | 291 (42%) |
| **Age at Accession** | **Median (IQR)** | 61 (52 - 70) | 63 (54 - 71) |
| **Anticoagulant**<br>**Subclass** | **Dabigatran** | 35 (0%) | 3 (0%) |
|  | **Heparinoid** | 135 (1%) | 11 (2%) |
|  | **None** | 10925 (96%) | 645 (95%) |
|  | **Vitamin K Antagonist** | 143 (1%) | 10 (1%) |
|  | **Xa-Direct Oral Anticoagulant** | 178 (2%) | 7 (1%) |
| **Metastatic Disease** | **No** | 3312 (28%) | 121 (17%) |
|  | **Yes** | 8383 (72%) | 572 (83%) |
| **Tumor Type** | **Bladder** | 400 (3%) | 32 (5%) |
|  | **Breast** | 1690 (14%) | 40 (6%) |
|  | **Colorectal** | 1084 (9%) | 62 (9%) |
|  | **Esophagogastric** | 286 (2%) | 30 (4%) |
|  | **Gyn** | 696 (6%) | 51 (7%) |
|  | **Head and neck** | 327 (3%) | 10 (1%) |
|  | **Hepatobiliary** | 383 (3%) | 32 (5%) |
|  | **High-grade glioma** | 487 (4%) | 44 (6%) |
|  | **Lung** | 1978 (17%) | 161 (23%) |
|  | **Melanoma** | 470 (4%) | 27 (4%) |
|  | **Other** | 1894 (16%) | 87 (13%) |
|  | **Pancreatic Adenocarcinoma** | 487 (4%) | 57 (8%) |
|  | **Prostate Adenocarcinoma** | 761 (7%) | 24 (3%) |
|  | **Renal** | 325 (3%) | 9 (1%) |
|  | **Soft Tissue Sarcoma** | 427 (4%) | 27 (4%) |
| **VTE Prior to Cancer**<br>**Diagnosis** | **No** | 11530 (99%) | 673 (97%) |
|  | **Yes** | 165 (1%) | 20 (3%) |

\* Cytotoxic chemotherapy anytime from day -30 to day 365 from accession date.

**Figure 1:**
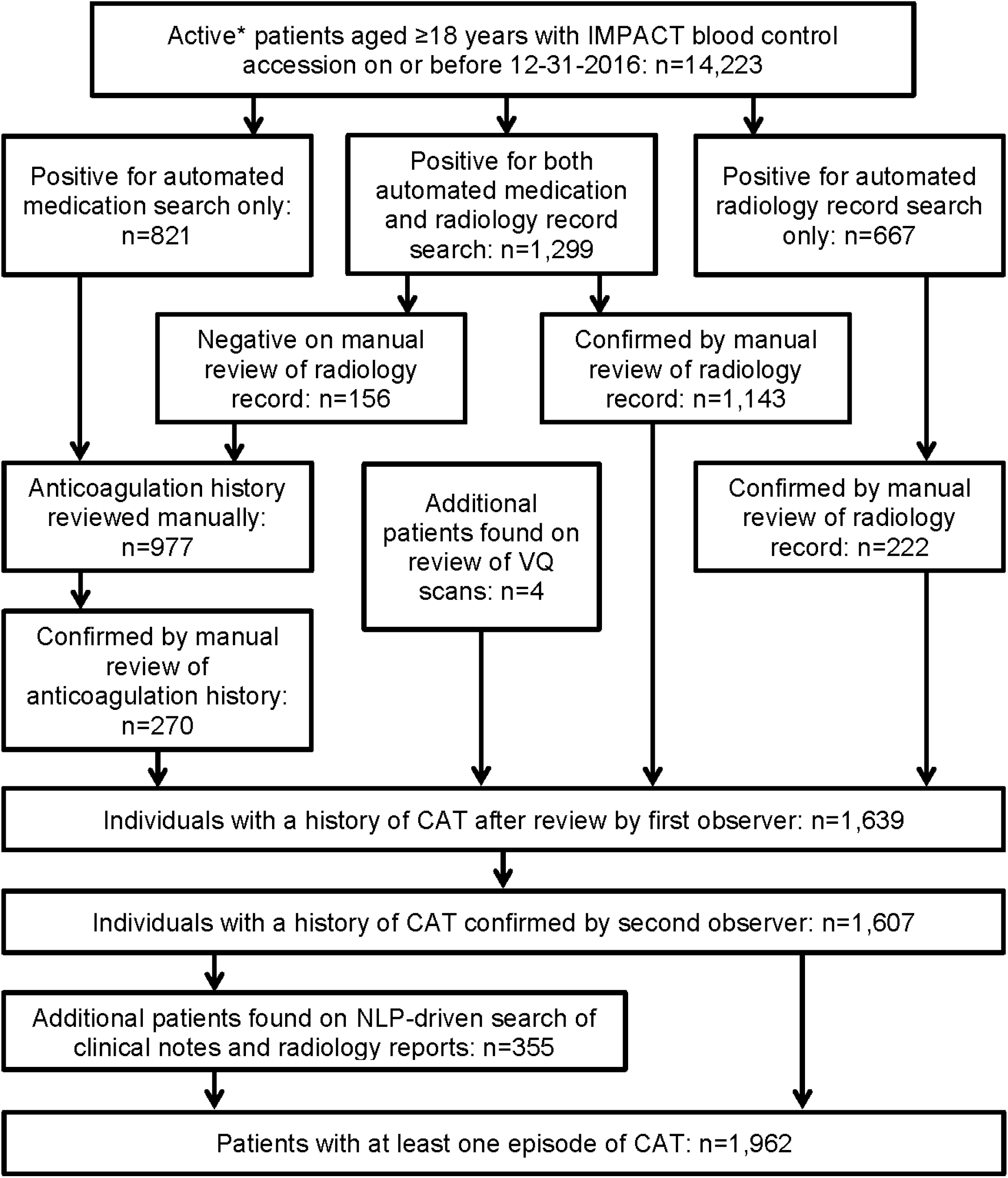
Flow of Cancer-Associated Venous Thromboembolism Event Assessment. *Patients who were followed actively at the medical center for the years 2014 to 2016. CAT: cancer-associated venous thromboembolism. NLP: natural language processing.

**Figure 2:**
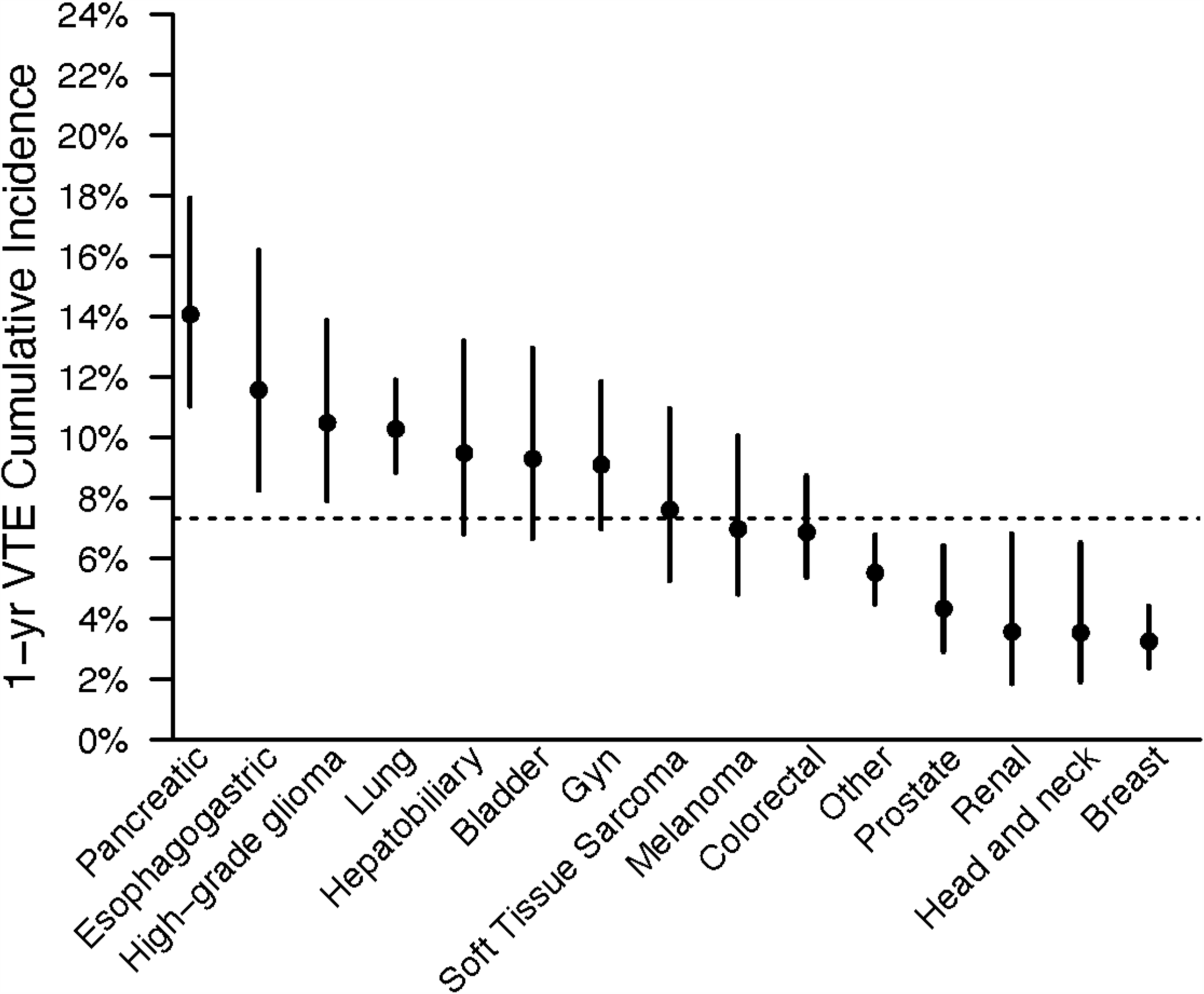
One-Year Incidence of Cancer-Associated Venous Thromboembolism by Tumor Type.

Clinical risk factors were assessed for VTE risk. Consistent with previous reports, cytotoxic chemotherapy (HR 1.63; 95% CI 1.32-1.93, *p*<0.001), prior VTE episode (HR 2.20; 95% CI 1.40-3.47, *p*=0.001), and metastatic disease (HR 2.60; 95% CI 2.03-3.33, *p*<0.001) were all strongly associated with CAT in multivariate analysis. Similarly, older age, an elevated white blood count, a decreased hemoglobin, an increased platelet count and higher BMI values were associated with a higher risk of CAT. Data on those latter parameters were not available for all patients.

### Specific somatic alterations in tumor are associated with increased venous thromboembolism risk

MSK-IMPACT data were then assessed to determine associations between individual genes and VTE risk across solid tumor malignancies. Tumor molecular profiles were analyzed across all cancer types and within sub-groups. Mutational frequencies across tumor types revealed rates of somatic alteration similar to those observed in previously published reports **(Supplementary Figure 1)**.^43^ *TP53* mutations had the highest prevalence, present in 42% of all tumor samples. *KRAS* and *EGFR* mutations were also frequent, found in up to 17% and 6% of tumor samples respectively and were identified across all cancer types, with a particularly high incidence in colorectal and pancreatic cancers. *IDH1* mutations were found to be enriched in glioma (24.9% of these patients), consistent with prior reports.^44^

There was no significant association between tumor mutational burden and risk of VTE (**Supplementary Figure 2**). There was no association of VTE risk with microsatellite instability (data not shown). Each gene was assessed in a separate regression model with the following covariates: age, previous VTE episode, anticoagulation, presence of metastatic disease and cytotoxic chemotherapy (time-dependent). Models were stratified based on tumor type and the time from a prior procedure to blood sample receipt for IMPACT germline control. Notably, mutations in *KRAS* (HR=1.34 [1.09-1.64]; adjusted *p*=0.08), *STK11* (HR=2.12 [1.55-2.89]; adjusted *p*<0.001), *KEAP1* (HR=1.84 [1.21-2.79]; adjusted *p*=0.07), *CTNNB1* (HR=1.73 [1.15-2.60]; adjusted *p*=0.09), *CDKN2B* (HR= 1.45 [1.13-1.85]; adjusted *p*=0.07), and *MET* (HR=1.83 [1.15-2.92]; adjusted *p*=0.09) were found to be associated with a significantly increased risk of CAT independent of tumor type (**Table 2**). Mutations in *SETD2* were associated with a decreased risk of CAT (HR=0.35 [0.16-0.79]; adjusted *p*=0.09). The unadjusted cumulative incidences of VTE for individuals with vs without a somatic mutation in these 7 genes are shown in **Figure 3**. Results stratified by tumor type are shown in **Supplementary Figure 3**. *CDK4* and *CDKN2A* mutations exhibited a trend towards an increased risk of CAT, with no statistically significant effect found after FDR-adjustment. A decreased risk of CAT was noted with *IDH1* mutations, but this effect was not significant after adjustment.

**Table 2:**
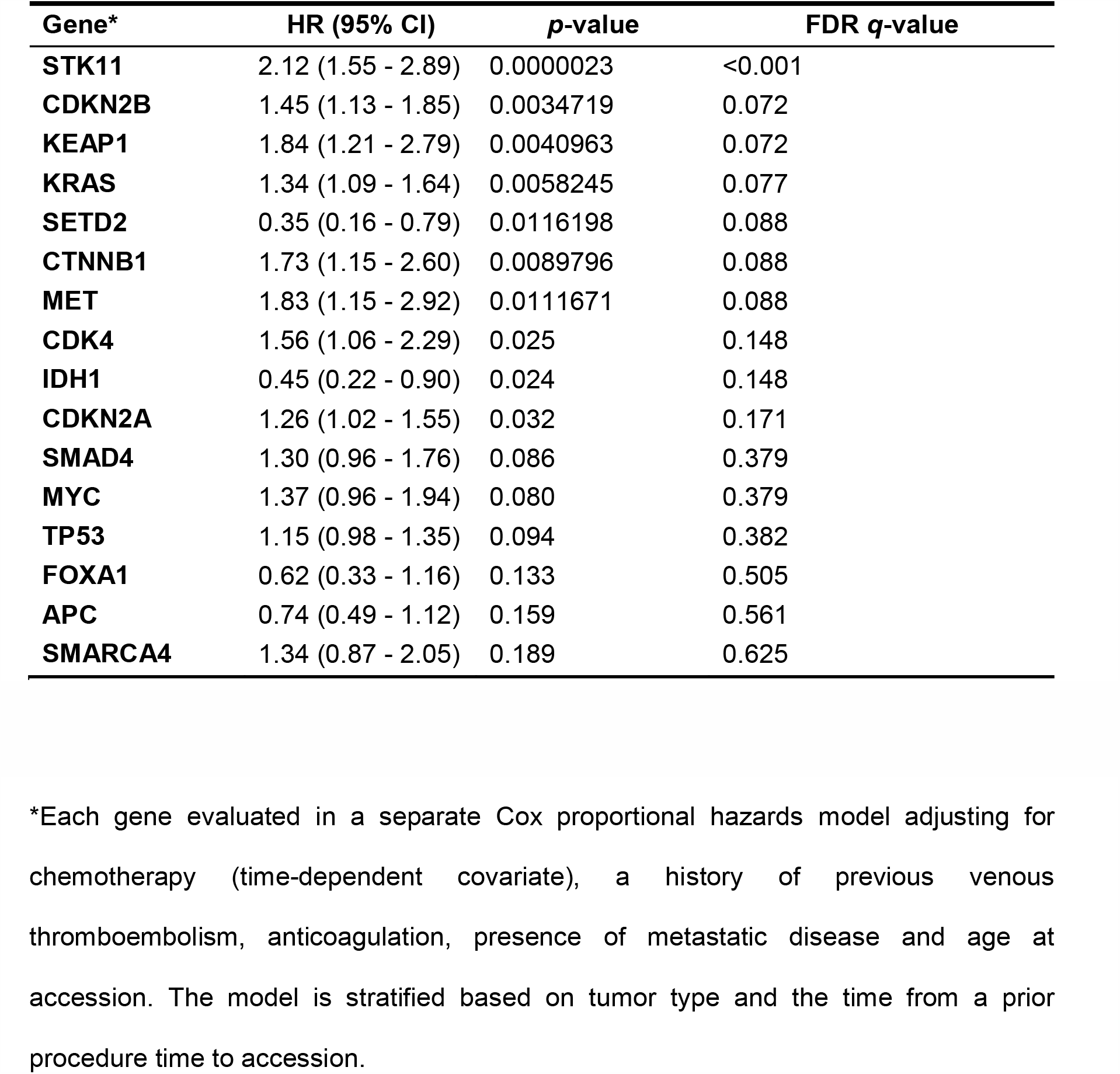
Hazard Ratio of Venous Thromboembolism for Selected Cancer Somatic Mutations.

**Figure 3:**
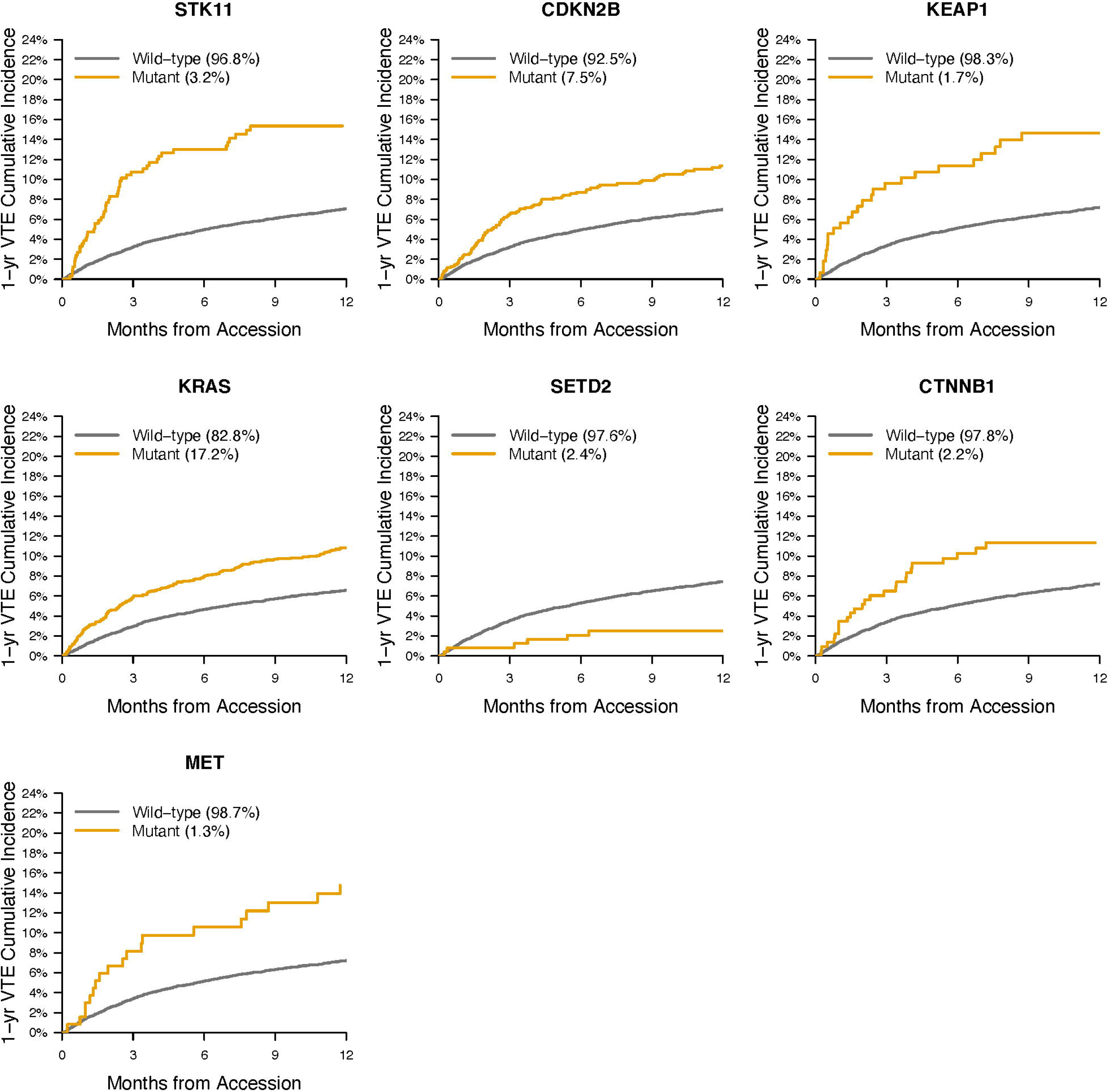
One-Year Cumulative Incidence of Cancer-Associated Venous Thromboembolism by Mutation Type.

Given the observed effect of *STK11* mutations on the risk of CAT, a dedicated analysis was conducted on data from The Cancer Genome Atlas (TCGA) to evaluate the effect of *STK11* mutations on RNA expression for tissue factor (*F3*) and G-CSF (*CSF3*). Tissue factor is widely believed to be an important effector of CAT and has been shown to be upregulated by several oncogenes.^34^ G-CSF has been demonstrated to be a likely mediator of the neutrophilia observed in lung cancer, also potentially being responsible for the formation of neutrophil extracellular traps and thromboembolism.^45,46^ A dataset including 503 patients from TCGA and with lung adenocarcinoma was selected in *cBioPortal*, given the expected high prevalence of *STK11* mutations in this group.^41,42,47^ RNA expression values were batch normalized (RNA Seq V2 RSEM) and a z-score threshold of +/- 2.0 was used. The mRNA expression z-scores relative to all samples were 0.58 vs −0.09 for *F3* (*STK11* mutated vs unmutated, *p*=1.334E-07) and 0.39 vs −0.15 for *CSF3* (*STK11* mutated vs unmutated, *p*=9.175E-05), as shown in **Supplementary Figure 4**. In order to assess the potential impact of increased transcription of G-CSF, a linear model predicting absolute neutrophil count based on the presence of a *STK11* mutation and adjusting for cancer type was fit using the MSK-IMPACT cohort. The presence of a *STK11* mutation was associated with an increase in 1,223 cells/mcL for the absolute neutrophil count (*p*<0.001).

### Clonal hematopoiesis in solid tumor patients does not increase the risk of VTE

We next assessed MSK-IMPACT data for the presence of CH and association with CAT. Of the 11,695 patients in the final cohort, 30% were found to have CH with a mutation in a known CH gene (**Supplementary Figure 5**). There was no significant association between any CH mutations, including *JAK2* V617F, and differential risk of CAT in the observed cohort (data not shown).

## Discussion

This work is the first broad search of a DNA tumor registry of its kind aimed to elucidate cancer-specific genomic determinants of CAT. Identifying mutations that might modify a patient’s risk for CAT might not only improve prognostication, but also potentially uncover new pathophysiologic mechanisms of how aberrant signaling might lead to VTE development. The results of this study identified multiple individual tumor mutations influencing VTE risk in solid malignancy patients, including *KRAS* for which prior data exist.^20,21,48^ Pre-clinical data demonstrate aberrant signaling through activating mutations of *MET* might confer a greater risk of CAT,^33,49^ and this study suggests this finding applies to human neoplasms. Additionally, *IDH1* mutations in gliomas have been demonstrated to be associated with a lower risk of VTE,^31,32^ a finding supported by our analysis even though the effect did not reach statistical significance after adjustment for multiple comparisons. On the other hand, the increased risk of CAT demonstrated with mutated *STK11, CDKN2B, CTNNB1* and *KEAP1* genes has not been previously reported. However, the effect of *KEAP1* was mitigated in multi-gene regression models (data not shown), and mutations in this gene are strongly correlated with *STK11* defects^50^ suggesting *KEAP1* mutations might not be an independent predictor of CAT risk. The mechanisms by which *STK11* mutations are associated with an increased risk of VTE are unclear, and might include increased neutrophil extracellular trap formation secondary to G-CSF production by the tumor. The possible explanations behind the decreased risk of CAT seen with *SETD2* mutations also remain to be determined.

Notably, previous studies revealed conflicting results in regard to *EGFR* mutational status in lung cancer and VTE risk, with either a protective effect, no effect or an increased risk.^20,25-28^ In this large retrospective cohort including most solid tumor types, we failed to establish a significant association between CAT risk and the presence of an *EGFR* mutation. One possible explanation would be that in the last few years, patients with *EGFR*-mutated lung cancer have received *EGFR*-targeted therapies. In this regard, anti-*EGFR* monoclonal antibodies have been associated with an increased risk of arterial and venous thromboembolism, while *EGFR*-targeted tyrosine kinase inhibitors have not.^51,52^ As such, differences in exposure to *EGFR*-directed therapies in different cohorts of patients with lung cancer could potentially result in discordant estimates of thromboembolism risk.

Available peripheral blood sequencing data for this large cancer cohort also allowed for the detection of CH and assessment of its effect on CAT risk. No significant association was found between the presence of any CH mutation and elevated VTE risk across all tumor types, including *JAK2* V617F, despite a previously published report of this association in non-cancer patient populations.^38^ Germline mutations were also excluded from this analysis.

Any retrospective cohort study such as this is prone to several potential pitfalls in terms of data collection and analysis. Ascertainment of clinical events on a cohort of more than 10,000 individuals is particularly challenging. In the present case, only a fraction of the available clinical notes were reviewed by a human observer so it is possible that a small number of CAT events were missed. In addition, another important aspect in determining the validity of results for a clinical genomic study is the method of classifying genetic data with respect to mutational status. We decided to use a simple approach, labeling individual cohort members as being mutated or not for any given gene. One might devise a more sophisticated method and sub-classify participants based on the specific mutations encountered for the most commonly mutated genes like *KRAS*. Estimation of the false discovery rate (FDR) is another very important aspect of cancer genomic studies, and we chose 0.10 as a cutoff for significance. It is very possible that some genes with an FDR >0.10 in our models might be significant predictors of CAT. In this regard, any analysis measuring multiple associations requires special care to interpret results of statistical significance tests. How these mutations might contribute to increased thrombosis development is not known and will require further validation along with functional studies.

## Conclusion

Improved risk stratification methods for VTE risk are needed for the diagnosis and prevention of CAT. Enhanced genetic tools allow for the detection of molecular events that might contribute to CAT risk. Retrospective research such as the work presented here can help elucidate the pathophysiology of CAT and aid in the development of risk stratification tools, however the high dimensionality of the datasets used is challenging. The analysis we performed on a large cohort of patients with solid tumors suggests associations between cancer-specific mutations and CAT risk, justifying further validation of key results along with dedicated functional studies to better elucidate how tumor-specific alterations contribute to thrombotic disease in this high-risk population.

## Data Availability

Genomic data is available publicly on cBioPortal. Other data available on request from.

http://cbioportal.org

## Acknowledgements

The results published here are in part based upon data generated by the TCGA Research Network: https://www.cancer.gov/tcga.

Memorial Sloan Kettering research support: NIH/NCI Cancer Center Support Grant P30 CA008748.

Dr. Dunbar receives support from the American Association of Cancer Research (17-40-11-DUNB)

Dr. Khorana acknowledges additional research support from the Sondra and Stephen Hardis Chair in Oncology Research and the National Heart, Lung and Blood Institute (U01HL143402).

Dr Park receives funding from the NIH (K12 CA184746 Paul Calabresi Award), the Parker Institute for Cancer Immunotherapy and SITC - Sparkathon TimIOs.

Dr Iyengar receives research support from the NCI/NIH (R01CA235711), the Breast Cancer Research Foundation and the American Cancer Society.

J.G. was supported by the Marie-Josée and Henry R. Kravis Center for Molecular Oncology.

## Authorship Contributions

S.M. planned the study and contributed to data collection, analysis and drafting of the manuscript.

A.D. drafted the manuscript and helped oversee the analysis.

S.M.D. performed the statistical analysis.

K.L.B. contributed to the analysis and drafting of the manuscript.

J.V.M., M.F., K.C. and Y.P. helped with data collection.

F.S.V. and J.G. contributed to the analysis.

K.J., J.W. and D.K. helped with data aggregation.

N.M.I. contributed to planning of the study.

R.L.L., S.K., A.Z., W.P., A.A.K. and G.A.S. contributed to drafting of the manuscript.

## Figure Legends

**Supplementary Figure 1:** Frequency of Mutations

**Supplementary Figure 2:** Cumulative Incidence of Cancer-Associated Venous Thromboembolism by Tumor Mutational Burden for Most Frequently Observed Tumors

**Supplementary Figure 3:** One-Year Cumulative Incidence of Cancer-Associated Venous Thromboembolism by Mutation Type Stratified by Tumor Type*

*Two tumor types with the highest mutation frequencies used for each gene

**Supplementary Figure 4:** mRNA Expression Z-scores Relative to All Samples for F3 (A) and CSF3 (B) for Patients With vs Without a STK11 Mutation*

*Data from TCGA Pan-Cancer Atlas, all samples are from adenocarcinoma of the lung, n=503.

**Supplementary Figure 5:** Cumulative Incidence of Cancer-Associated Venous Thromboembolism by Clonal Hematopoiesis Status

PanCan PD: pan-cancer, putative driver clonal hematopoiesis CH: clonal hematopoiesis

